# Development of a novel transcriptomic measure of aging: Transcriptomic Mortality-risk Age (TraMA)

**DOI:** 10.1101/2024.12.04.24318517

**Authors:** Eric T. Klopack, Gokul Seshadri, Thalida Em Arpawong, Steve Cole, Bharat Thyagarajan, Eileen M. Crimmins

## Abstract

Increasingly, research suggests that aging is a coordinated multi-system decline in functioning that occurs at multiple biological levels. We developed and validated a transcriptomic (RNA-based) aging measure we call Transcriptomic Mortality-risk Age (TraMA) using RNA-seq data from the 2016 Health and Retirement Study using elastic net Cox regression analyses to predict 4-year mortality hazard. In a holdout test sample, TraMA was associated with earlier mortality, more chronic conditions, poorer cognitive functioning, and more limitations in activities of daily living. TraMA was also externally validated in the Long Life Family Study and several publicly available datasets. Results suggest that TraMA is a robust, portable RNAseq-based aging measure that is comparable, but independent from past biological aging measures (e.g., GrimAge). TraMA is likely to be of particular value to researchers interested in understanding the biological processes underlying health and aging, and for social, psychological, epidemiological, and demographic studies of health and aging.

## Introduction

A growing body of research suggests that aging is a coordinated multi-system decline in functioning that occurs at multiple biological levels (e.g., DNA damage accumulation, cellular aging and senescence, chronic disease morbidity, physical disability)^1,2^. A major goal of geroscience research is to develop biomarkers of this aging process using minimally invasive methods in humans, as these markers are highly useful in evaluating interventions, understanding social inequalities in health and aging, and researching causes and consequences of accelerated aging in humans^3,4^. Biomarkers of aging have been developed using combinations of clinical biomarkers^5,6^, DNA methylation^3,7–10^, inflammatory markers^11^, telomere length^12^, metabolomics^13,14^, and proteomics^15,16^. These tools have been extremely useful for understanding how social and environmental exposures affect health and aging^17–23^, the long-term impact of early life adversity^24–27^, how timing of exposure matters for health^28–30^, among other important advances.

Previous transcriptomic (RNA-based) aging measures^31^ were generally developed using array data (rather than RNA sequencing, which predominate in newer studies), have utilized small, specialty samples, or were estimated in tissue other than blood^31–33^. Indeed, a recent review noted the limitations of existing transcriptomic aging measures and the large number of unknowns about their reproducibility and ability to capture health and mortality risk^34^. At the same time, there has been a proliferation of research utilizing next-generation high throughput RNA sequencing (RNAseq), and several large population-based surveys (e.g., the Health and Retirement Study (HRS), the National Longitudinal Study of Adolescent to Adult Health (Add Health), Midlife in the United States (MIDUS), the Northern Ireland Cohort for the Longitudinal Study of Ageing (NICOLA)) are collecting large RNAseq samples that will be able to address questions about the causes and consequences of transcriptomic aging at the population level.

For these analyses to yield useful generalizable findings, a reliable and portable summary measure of accelerated transcriptomic aging is needed. We developed such a measure here using the 2016 HRS Venous Blood Study (VBS), a nationally representative sample of nearly 4000 US adults aged 50 and older. We utilized elastic net penalized regression to estimate a transcriptomic prediction measure of 4-year mortality risk—Transcriptomic Mortality-risk Age (TraMA)—using more than 10,000 gene transcripts in a training sub-sample. We evaluated this measure in a hold-out testing sub-sample of the HRS, in an external dataset (the Long Life Family Study; LLFS), and in several other publicly available datasets. Our plan of analysis for this study is shown in Figure 1, Panel A.

**Figure 1.**
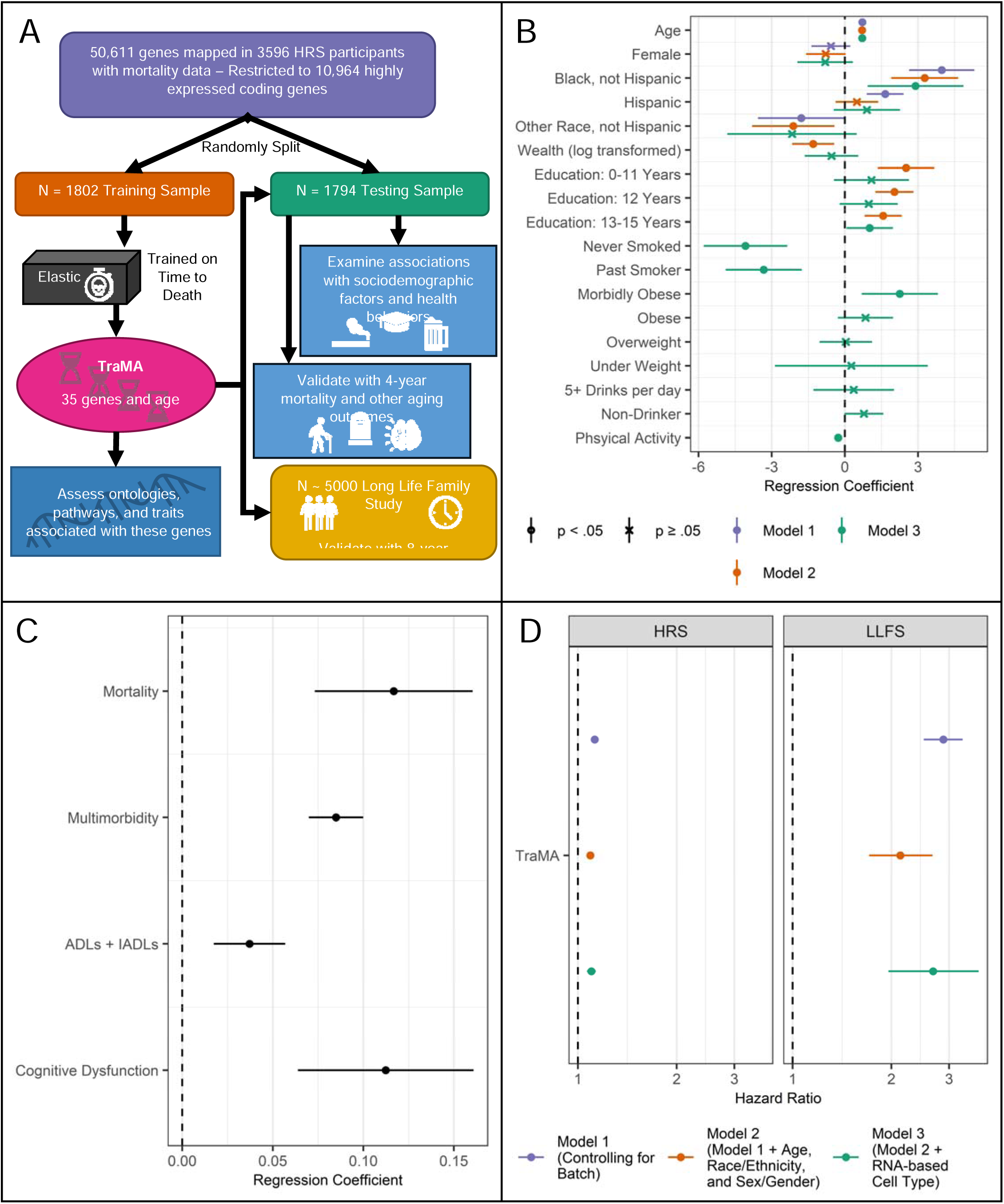
***Panel A.*** Plan of analysis for the current study. ***Panel B.*** Nested regression results from the HRS testing data including associations between TraMA and sociodemographic factors and health behaviors; points represent regression coefficients and bars represent 95% confidence intervals; all models include cell type and batch as covariates. Model 1 includes demographic factors; Model 2 includes variables in Model 1, as well as socioeconomic factors; Model 3 includes variables in Model 2, as well as health behaviors. ***Panel C.*** Regression results from the HRS testing data of health/aging outcomes on TraMA; points represent regression coefficients and bars represent 95% confidence intervals; all models include age, race/ethnicity, sex/gender, cell type, and batch as covariates. ***Panel D.*** Validation results from nested regression of time to death on TraMA in HRS and LLFS. Model 1 includes batch as a covariate; Model 2 includes batch, age, race/ethnicity, and sex/gender as covariates; Model 3 includes variables from Model 2, as well as RNA-based cell type as covariates.

## Results

### Training TraMA

Because we are interested in developing a measure that is accurate and portable to other human datasets, we restricted the set of genes used for training to coding genes with relatively high expression in human venous blood. To accomplish this, of the 50,611 transcripts that were measured and were successfully mapped in HRS, we restricted ourselves to the 19,291 protein coding genes. We further restricted ourselves to genes with a mean count per million greater than 3 in the total HRS sample, leaving 10,964 genes. We also included chronological age in years and sex/gender as features to reduce age and sex/gender bias in the selection of genes (i.e., they are covariates along with the 10,964 genes used to train TraMA).

We then randomly split the HRS sample into a training (N = 1801) and testing (N = 1794) subsamples. Descriptive statistics for each subsample are shown in Table 1. 222 participants in the training data died during the 4-year follow-up. 197 participants in the testing data died during follow-up. We ran elastic net models predicting 4-year mortality in the training sample. Hyperparameters, including the alpha and lambda penalty terms, were selected using a grid search procedure with 5x cross validation. Log_2_ adjusted counts per million (log_2_cpm) were used in all analyses, including the Health and Retirement Study (HRS), the Long Life Family Study (LLFS), and publicly available data described below. This procedure selected an alpha of 1 (equivalent to LASSO regression) and a lambda of 0.0198. This model selected (i.e., did not reduce regression coefficients to 0) 35 genes and age. Gene names, Ensembl IDs, and coefficients are shown in Table 2 [HIDDEN BEFORE PEER REVIEW]. TraMA scores were transformed to have a mean and variance equivalent to the HRS training set chronological age. In the training data, Harrel’s C-index predicting survival from the TraMA score was 0.835, suggesting very good fit.

**Table 1a.** Weighted Descriptive Statistics for the HRS Training and Testing.

|  | Training Data |  | Testing Data |  |
| --- | --- | --- | --- | --- |
|  | Mean /<br>Proportion | SD | Mean /<br>Proportion | SD |
| Age | 68.62 | 9.21 | 68.66 | 9.09 |
| Race: Non-Hispanic White | 0.78 |  | 0.78 |  |
| Race: Non-Hispanic Black | 0.10 |  | 0.11 |  |
| Race: Hispanic | 0.08 |  | 0.09 |  |
| Race: Non-Hispanic Other Race | 0.04 |  | 0.02 |  |
| Gender: Female | 0.55 |  | 0.54 |  |
| Mortality | 0.11 |  | 0.10 |  |
| Multimorbidity | 2.12 | 1.39 | 2.04 | 1.37 |
| Cognitive Dysfunction | 11.56 | 4.31 | 11.31 | 4.35 |
| ADLs + IADLs | 0.72 | 1.74 | 0.64 | 1.60 |

### Gene Ontologies, Associated Traits, Pathway Analysis, and Functional Enrichment Analysis

#### Gene ontologies

A major value of biological aging measures is that they describe biological aging pathways, marking processes that underly health and aging that may not be phenotypic yet. That is, these measures help assess pre-diagnostic states before morbidities and mortalities manifest. To assess how well TraMA indexes these pathways, we assessed ontologies provided by Ensembl^35^. According to these ontologies, there is evidence several of these genes are involved in neurological development and functioning (*CNTNAP2*, *KCNA2*, *KIFBP*, *NELL2*, *NOG*), amyloid formation and regulation (*ADAM17*, *APH1B*), immune responses (*ADAM17*, *CLEC4C*, *GPR15*, *MCOLN2*), cell cycle regulation (*CDKN2B*, *TRIM39*), and methylation and gene expression regulation (*METTL9*, *ZNF417*, *ZNF44*). These ontologies, thus, include pathways essential to aging and health.

#### Associated traits

We assessed associated traits from past transcriptome-wide association studies (TWAS) for the 35 genes using TWAS logged in the TWAS Atlas^36^. A large number of these genes have been linked in past TWAS to body height (*ABTB3*, *ADAM17*, *ANGPT1*, *DSP*, *HDGFL3*, *KIFBP*, *LASP1NB*, *NKD1*, *PLVAP*, *TMEM38A*, *ZNF417*, *ZNF44*), weight and BMI (*ABTB3*, *ADAM17*, *ANGPT1*, *C12orf76*, *HDGFL3*, *KIFBP*, *NOG*, *PLVAP*, *SLC4A10*), blood pressure and hypertension (*C12orf76*, *CTTNBP2NL*, *SLC16A1*, *SLC4A10*, *TRIM39*), lung functioning (*DSP*, *LASP1NB*) and to chronological age (*ADAM17*, *CDKN2B*, *METTL9*). Thus, these genes have been associated with a number of age and development-related traits in past research.

#### Pathway and Functional Enrichment Analysis

We performed functional enrichment analysis using GeneMANIA. This program identifies gene ontology terms enriched among the list of genes identified in pathways and provides FDR corrected Q values and coverage ratios^37^. A large number of these functions involve the renal system, including nephron development, glomerulus development, kidney vasculature development, kidney development, renal system vasculature development, and renal system development. A number are also related to basic cell functioning (cell adhesion mediator activity, regulation of transmembrane receptor protein serine/threonine kinase signaling pathway, transmembrane receptor protein serine/threonine kinase signaling pathway) and cell cycle regulation (regulation of pathway-restricted SMAD protein phosphorylation, pathway-restricted SMAD protein phosphorylation). Other functions include nervous system (main axon, neuron recognition) and other biological system functioning. These functions make sense given the essential functions of the kidney and nervous system in aging and mortality and given the importance of cell cycle regulation for cellular aging.

### Validation in the Health and Retirement Study Testing Sample

#### Validation with time to death

We tested this surrogate score in the N = 1794 HRS hold-out testing sample. TraMA was significantly associated with mortality hazard in the testing subsample with age, sex/gender, race/ethnicity, and batch as covariates (HR = 1.09, 95% CI = [1.06, 1.12], p < 0.0001). Thus, having a TraMA score 10 years older (a little more than a standard deviation), was associated with about a 90% increase in mortality hazard. Harrel’s C index (an extension of area under the curve for survival data) was 0.81 indicating excellent fit.

#### Associations with sociodemographic factors and health behaviors

Researchers using measures of biological aging (e.g., epigenetic clocks, telomeres) are often interested in understanding how psychosocial, demographic, and behavioral risk factors contribute to differences in mortality and other health and aging outcomes. To indicate the utility of TraMA to these researchers we conducted a series of nested regressions, first regressing TraMA on basic demographic factors (viz., age, race/ethnicity, and sex/gender), then on socioeconomic factors thought to contribute to demographic differences in health (viz., wealth and education), and finally on health behaviors thought to mediate these sociodemographic associations with health (viz., smoking status, BMI (as a proxy for diet and activity), alcohol use, and a physical activity index. All regressions included RNA-based cell type distribution in whole blood (using log_2_cpm for *CD3D*, *CD19*, *CD4*, *CD8A*, *FCGR3A*, *NCAM1*, and *CD14*) and batch as covariates. Results are shown in Figure 1, Panel B.

In all models, chronological age is significantly associated with TraMA, which is expected as age is used in the calculation of TraMA. Compared to non-Hispanic White respondents, non-Hispanic Black respondents had a significantly higher TraMA in all models; Hispanic participants had an older TraMA without controlling for socioeconomic status or health behaviors; and non-Hispanic participants from other racial groups had a lower TraMA in models without health behaviors as covariates. Before controlling for health behaviors, greater wealth and higher educational attainment were associated with younger TraMA. After including health behaviors as covariates, having 13-15 years of education (vs 16 or more) was associated with older TraMA. Compared to current smokers, never smokers and past smokers had younger TraMA. Compared to normal weight individuals, morbidly obese (BMI ≥ 35) had older TraMA. Greater physical activity was associated with younger TraMA.

#### Associations with mortality and other health/aging outcomes

To assess TraMA as a general measure of aging, we also regressed multimorbidity (count of diagnoses with high blood pressure, diabetes, cancer, lung disease, heart disease, stroke, and arthritis), 4-year mortality, cognitive dysfunction (using errors on the Telephone Interview for Cognitive Status (TICS)), and count of at least some difficulties with activities of daily living (ADLs) and instrumental activities of daily living (IADLs), controlling for chronological age, race/ethnicity, sex/gender, RNA-based cell type, and batch effects. Regression coefficients are shown in Figure 1, Panel C. TraMA was significantly associated with each of these outcomes in the HRS testing data.

#### Comparison with Other Biological Aging Measures

As noted above, a large number of biological aging measures have been produced using omics data, telomeres, and indices of blood-based biomarkers. To be a useful and innovative measure of aging, TraMA should 1) be associated with health outcomes to a similar or greater magnitude compared to these measures and 2) should be associated with health outcomes above and beyond these measures.

To assess this first point we regressed health and aging outcomes on TraMA, GrimAge (an epigenetic aging measure), PhenoAge (an epigenetic aging measure), and ExpandedAge (an index of biomarkers associated with aging) controlling for age, race/ethnicity, sex/gender, RNA-based cell type, and batch, shown in Figure S1, Panel A. Each point in this panel represents results from a separate regression. Associations between TraMA and health outcomes are slightly stronger than those of PhenoAge or ExpandedAge. Association between TraMA and mortality and between TraMA and cognitive dysfunction are slightly weaker than similar associations with GrimAge. TraMA has a very similar, but slightly higher, association with multimorbidity compared to GrimAge. TraMA, PhenoAge, and ExpandedAge, but not GrimAge, were significantly associated with ADLs and IADLs. Thus, the pattern of significant results was highly consistent across these aging measures.

Given these similarities, one may wonder whether TraMA explains unique variance in health outcomes or if it simply duplicates other existing aging measures. To address this issue, we regressed each health outcome on TraMA and GrimAge in the same model, along with age, race/ethnicity, sex/gender, RNA-based cell type, and batch, shown in Figure S1, Panel B. Both TraMA and GrimAge were significantly associated with mortality at very similar magnitudes. When in the same model together, TraMA, but not GrimAge, was associated with multimorbidity and ADLs and IADLs. When in the same model together, GrimAge, but not TraMA, was associated with cognitive dysfunction, though the association with TraMA approached significance (*p* = 0.066). Thus, TraMA appears to mostly describe unique variance in mortality, multimorbidity, and ADLs and IADLs compared to GrimAge.

### Validation in the Long Life Family Study

To ensure portability of this measure, and as a further check for robustness, we also validated this measure in an external cohort that includes a large number of older adult humans, the LLFS. Using mixed effect Cox proportional hazards models, we regressed time to death in each sample on TraMA, controlling for batch (Model 1); adding age, race/ethnicity (in HRS; because nearly all LLFS participants were White, we did not control for Race/Ethnicity in LLFS), and sex/gender (Model 2); and adding RNA-based cell type (Model 3). All models were adjusted for family relatedness as a fixed effect. Results are shown in Figure 1, Panel D. TraMA was significantly associated with time to death in both LLFS and HRS with all controls included.

The hazard ratio for LLFS is higher than for HRS. This potential because LLFS participants were selected for their longevity. LLFS also had twice the number of mortality events as HRS. Because the LLFS includes a sample of older adults, their children, and spousal controls, we additionally ran analyses only in the sample of older adults (referred to in the LLFS as the proband generation). These results largely replicated those shown in Model 3 above, with a hazard ratio of 2.48 and *p*-value less than 0.001 for the proband generation, 4.76 and *p*-value less than 0.01 for the offspring generation, and 1.63 and *p*-value less than 0.01 for the spousal controls.

### Validation in Small and Clinical Samples

To be a maximally valuable measure of the aging process, TraMA should be useful not only in large representative samples, but also in small specialty and clinical samples. We therefore additionally validated in four publicly available datasets from the Gene Expression Omnibus (GEO) with RNA-seq data from whole blood and information about chronological age. First, to validate expected associations with health behaviors, we estimated TraMA in data from 454 current and 767 former smokers from the COPDGene Study (GEO series GSE171730), including non-Hispanic White and African American people between the ages of 45 and 80 in the US (shown in Figure 2, Panel A)^38,39^. In these data, TraMA was positively associated with smoker status and number of smoking pack years, and negatively associated with lung functioning assessed with forced expiratory volume in one second (FEV1) predicted percentage. Current smokers had an estimated TraMA 2.83 years older than former smokers controlling for age, race, sex/gender, RNA-based cell type, and batch.

**Figure 2.**
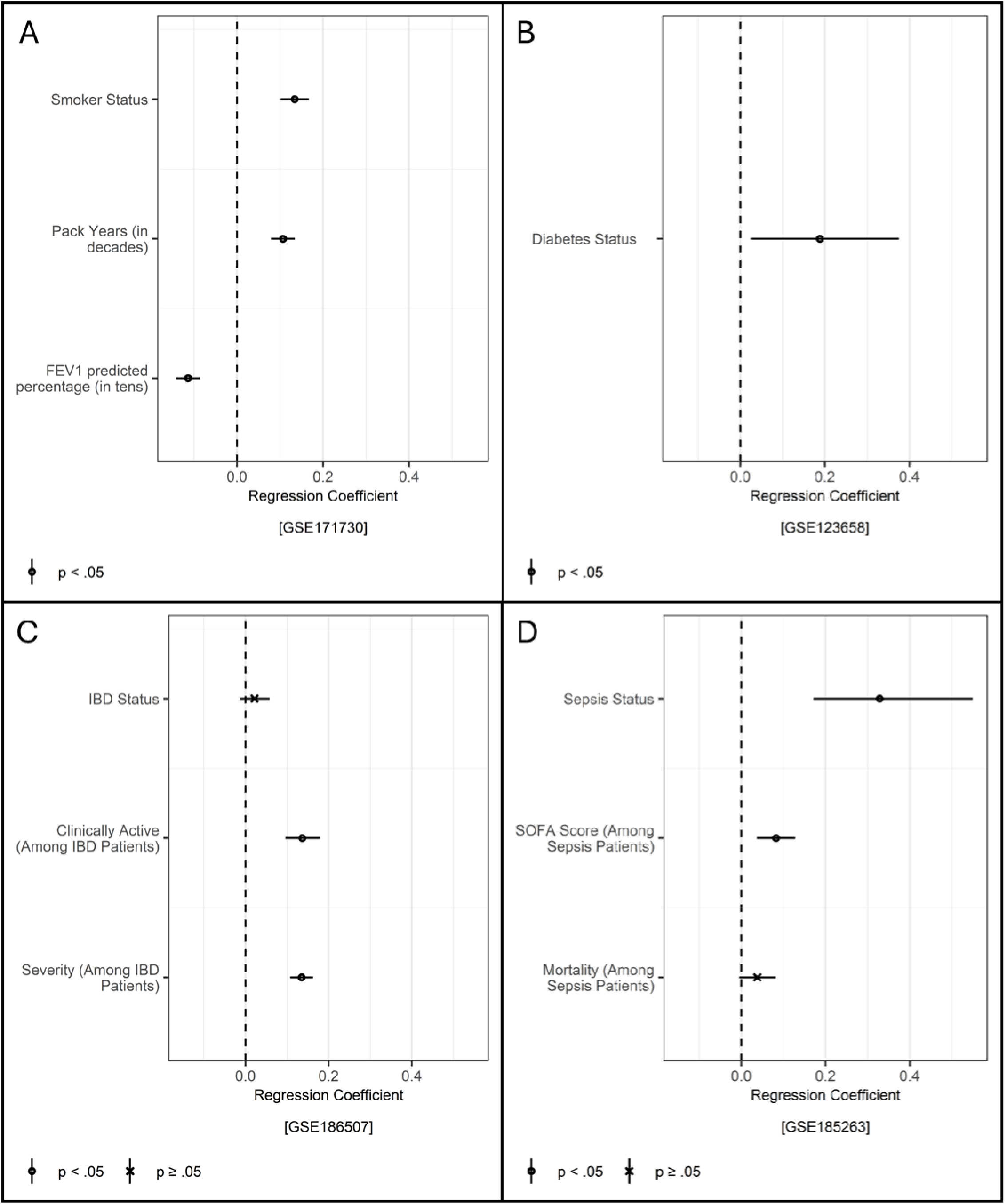
***Panel A.*** Regressions of smoker status, cigarette pack years (divided by 10), and forced expiratory volume over one second (FEV1) predicted percentage (divided by ten) on TraMA controlling for age, race, sex/gender, cell type, and batch; points represent regression coefficients and bars represent 95% confidence intervals. ***Panel B.*** Regression of diabetes status on TraMA controlling for age, sex/gender, and cell type; points represent regression coefficients and bars represent 95% confidence intervals. ***Panel C.*** Regressions of inflammatory bowel disease (IBD) status, clinically active IBD, and IBD severity from endoscopy on TraMA controlling for age, sex/gender, and cell type; points represent regression coefficients and bars represent 95% confidence intervals. ***Panel D.*** Regressions of sepsis status, sequential organ failure assessment (SOFA) score, and mortality among sepsis patients on TraMA controlling for age, sex/gender, and cell type; points represent regression coefficients and bars represent 95% confidence intervals.

TraMA was also associated with diabetes status in a sample of 43 healthy participants and 39 participants with type 1 diabetes (GEO series GSE123658; results shown in Figure 2, Panel B)^40^. In these data, controlling for age, sex/gender, RNA-based cell type, and batch, participants with diabetes had a predicted TraMA 2.12 years older than healthy controls.

We also validated in data from the Mount Sinai Crohn’s and Colitis Registry (MSCCR; GEO series GSE186507)^41^ including 821 participants with inflammatory bowel disease (IBD; 432 with Crohn’s disease and 389 with ulcerative colitis) and 209 healthy controls (results shown in Figure 2, Panel C). IBD status was not significantly associated with TraMA after statistically controlling for age, sex/gender, and RNA-based cell type; however, this association was significant without RNA-based cell type (log odds = 0.07, *p* < .001), and Crohn’s disease participants had elevated TraMA compared to health controls with all covariates (b = 1.00, *p* < .05), this may be because Crohn’s disease patients were more likely to have active IBD (Harvey-Bradshaw index (HBI) ≥ 5; *p* < .01). Among all IBD patients, participants with clinically active IBD (according to a physician’s evaluation) had higher TraMA, and TraMA was positively associated with IBD severity using the Simple Endoscopic Score for Crohn’s Disease (SESCD).

TraMA was associated with sepsis, compared to healthy controls, in a sample of 348 sepsis patients and 44 healthy controls (GEO series GSE171730; results shown in Figure 2, Panel D)^42^ in a regression including age, sex/gender, and RNA-based cell type as covariates. Compared to healthy controls, sepsis patients had an estimated TraMA 10.31 years older. TraMA was also associated with Sequential Organ Failure Assessment (SOFA) scores among sepsis patients. The association between TraMA and mortality among sepsis patients approached significance (*p* = 0.09), though this association was significant in a model without cell type (log odds = 0.05, *p* < .001).

## Discussion

Large, population-based studies of aging are collecting omic-level biological data, creating a unique and exciting opportunity to understand both population-level and potentially individual-level biological aging processes. Measures of aging have been developed using DNAm and sets of clinical and blood-based biomarkers, and these measures have rapidly advanced research on aging and health. However, a similar measure does not exist for RNAseq data. This is a major gap in past research, as RNA represents a critical step in gene expression and, ultimately, nearly all biological processes. To address this gap, we developed Transcriptomic Mortality-risk Age (TraMA) using RNAseq data in the HRS and validated this measure in the LLFS and in several publicly available datasets.

We used an elastic net approach to identify genes associated with 4-year all-cause mortality. This method is in line with so-called second-generation DNAm-based epigenetic clocks (e.g., PhenoAge^3^, GrimAge^9^) that focus on phenotypic indicators of aging as criterion variables. These second-generation clocks have been found to be much more strongly associated with both health outcomes and health risk exposures compared with first-generation clocks that used chronological age alone as a criterion variable^21,25^.

Thus, TraMA is in line with similar DNAm-based measures that have been consistently associated with health outcomes, health risk exposures, and sociodemographic factors^43,44^. Indeed, analyses here indicate that TraMA is similarly associated with health outcomes, including mortality, multimorbidity, ADLs and IADLs, and cognitive functioning. However, these analyses also find TraMA captures unique variance in age-related health outcomes, compared to GrimAge.

Genes selected by this elastic net procedure represented a number of developmental and health processes we would expect to be associated with aging and mortality (e.g., immune response, cell cycle regulation, gene expression regulation, body weight, blood pressure, and chronological age). Thus, this measure captures biologically plausible cellular and multi-system processes involved in aging.

This study is not without limitations. The HRS is representative of the older US population, but the aging process likely differs across national and cultural context. Because we were interested in assessing mortality risk as an indicator of aging in older adults, we utilized data from the HRS, where participants were all aged 50 or older. However, we validated this measure in samples that included relatively young participants (as young as 19).

Blood-biomarker-based and DNAm-based aging measures have rapidly accelerated aging research. We believe our RNA-based measure has the capacity to contribute to this highly active and quickly evolving literature. Associations between TraMA and health outcomes were robust and consistent in the HRS testing sample, the LLFS, and other validation samples. Thus, this measure appears to be a useful, portable indicator of the aging process. It appears to explain a large, unique portion of aging-related health outcomes and is associated with health risks in expected directions. Our results show its utility in both large, population-based sample, and smaller clinical, specialty, and community-based samples. We, thus, believe this measure can be a useful tool for researchers interested in understanding the aging process in humans.

## Online Methods

### Cohorts

***The Health and Retirement Study (HRS)*** is an ongoing panel study of older adults since 1992 that is designed to be representative of older US adults when weighted. As part of 2016 data collection, venous blood was collected from a subsample of the HRS. 2.5 ml of blood was collected in PAXgene tubes from about 4000 participants. Total RNA extraction was performed on the QIACube semi-automated method using the PAXgene Blood miRNA Kit. Assays used 200-500 ng of RNA for each sample. All RNA species were extracted and stored for future use. RNA was extracted from only half a PAXgene tube to ensure RNA storage in a variety of formats. Ribosomal RNA and globin reduction performed using the TruSeq stranded Total Library Prep Gold kit - Ribozero Gold kit. RNAseq was performed on a NovaSeq (Illumina Inc.) using 50 bp paired end reads. All samples were sequenced to a minimum depth of 20 M reads. RNA-Seq was successfully performed on 3685 participants. The HRS pipeline closely mirrors the TOPMed/GTEX RNA-Seq analysis pipeline with minor modifications. More information about RNAseq pipelines are available elsewhere^45^ including the HRS website (https://hrs.isr.umich.edu/about).

***The Long Life Family Study (LLFS)*** is a longitudinal sample of nearly 5000 participants from 539 families that were selected because of their exceptional longevity. There have been three waves of data collected 6-8 years apart. The first and second waves of data included blood collection. We use data from the first wave to align with HRS. More information is available at the LLFS website https://longlifefamilystudy.com/.

RNA sequencing for Visit 1 was performed using RNA extracted from PAXgene™ Blood RNA tubes, processed with the Qiagen PreAnalytiX PAXgene Blood miRNA Kit. Library preparation, quality control, and sequencing were carried out by the Division of Computation & Data Sciences at Washington University, using the nf-core/rnaseq 3.14.0 pipeline for read alignment, duplicate marking, and transcript quantification. Genes with low expression (fewer than 4 counts per million in at least 98.5% of samples) and those with significant intergenic overlap were filtered out. This resulted in a final dataset of 1,810 samples and 16,418 genes. For this study, we utilized RNAseq data from the LLFS dataset, with the filtered raw counts converted to a Log_2_CPM (counts per millsion) scale for further analysis.

***The COPDGene study (GSE171730)*** that is publicly available includes 454 current and 767 former smokers, including non-Hispanic White and African American men and women between the ages of 47 and 86 in the US. RNAseq was performed on whole blood using the Illumina HiSeq 2000 platform. More information is available on the COPDGene website (https://copdgene.org/). Information about current smoker status, pack years, and forced expiratory volume in one second (FEV1) predicted percentage, race, sex/gender, and batch are available.

***GSE123658*** is a sample of 43 healthy donors and 39 type 1 diabetes patients between ages 19 and 73. RNAseq was assessed in whole blood using Illumina NextSeq 500 or HiSeq 4000 platforms. Information about diabetes status, age, and sex/gender are available.

***The Mount Sinai Crohn’s and Colitis Registry (GSE186507)*** includes 821 irritable bowel disease (IBD) patients and 209 healthy controls aged 19 to 82 recruited during an endoscopy appointment form December 2013 to September 2016. RNAseq was assessed in whole blood using the Illumina HiSeq 2500 platform. Information about IBD status, active IBD status (Harvey-Bradshaw index (HBI) ≥ 5), disease severity Simple Endoscopic Score for Crohn’s Disease (SESCD), age, and sex/gender are available.

***GSE185263*** is a sample of 348 sepsis patients and 44 healthy controls aged 18 to 96 from countries, including Australia, Colombia, the Netherlands, and Canada (sites in Toronto and Vancouver). RNAseq was assessed in whole blood using the Illumina HiSeq 2500 platform. Information about sepsis status, severity using Sequential Organ Failure Assessment (SOFA) scores, mortality, age, and sex/gender are available.

### Measures

***Time to death in the HRS*** was assessed using information about date of interview and date of death from the HRS tracker file. We use 4-year mortality with participants know to be deceased to the HRS. Time to death was calculated as the difference between the 2016 interview month and the known month of death. For participants who survived (i.e., were not known to have died), time at risk was calculated as the time between the 2016 interview month and the most recent interview month available. This measure was used in elastic net models.

***Mortality in the HRS for logistic regression.*** We created a binary indicator of 4-year mortality used in logistic regression models.

***Time to death in the LLFS.*** Because relatively few of the LLFS participants died in 4 years compared to HRS, we used 8-year mortality for the replication analysis using participants known to be deceased by the LLFS. Time to death was calculated as the difference between 2006 blood sample collection date and the known date of death censored at 31 December 2014 (about 8 years after Wave 1 data collection).

***Multimorbidity*** was calculated in HRS as the sum of diseases that a participant had ever been told by a doctor that they had, including high blood pressure, diabetes, cancer, lung disease, heart disease, stroke, and arthritis.

***Cognitive dysfunction*** was assessed in HRS using the Telephone Interview for Cognitive Status (TICS). To make this a measure of dysfunction, we used errors (27 minus the sum of these scores) in immediate recall (10 words), delayed recall (the same 10 words after about 5 minutes of other survey questions), serial 7s (participants were asked to subtract 7 from 100 and continue subtracting for 5 trials), and backwards counting from 20 (participants were asked to count backward from 20 to 10 and given 2 points for a correct first try and 1 for a correct second try).

***ADLs and IADLs*** in HRS is the sum of self-reported difficulty with walking across a room, dressing, bathing, eating, getting in and out of bed, using the toilet, using a map, using the phone, taking medications, managing money, shopping for groceries, and preparing a meal.

***Other biological aging measures*** used in the current study include two epigenetic aging measures produced by HRS^46^, PhenoAge^3^ and GrimAge^9^, and Expanded Biological Age (ExpandedAge)^6^. PhenoAge and GrimAge are both so-called second-generation epigenetic clocks. They are indices of cytosine-phosphate-guanine (CpG) sites where differential methylation is associated with age-related biomarkers and mortality. These have been widely used in past research and have been of extraordinary value in advancing geroscience and in clarifying the biological processes underlying social, psychological, and demographic differences in health and aging^44,47–49^. We use so-called principle component versions of these clocks which have been shown to be more reliable^50^. ExpandedAge is an index of 22 biomarker of phenotypic aging that has been linked to mortality and other health outcomes^6^.

***Demographic factors*** used in regression analyses include chronological age in years, sex/gender (female as the reference group), and race/ethnicity (non-Hispanic Black, Hispanic, non-Hispanic other race, and non-Hispanic White as the reference group).

***Socioeconomic factors*** include years of education as reported to HRS (0-11 years, 12 years (the typical number of years for a high school degree in the US), 13-15 years, and 16 or more years as the reference group), as well as total wealth as calculated by RAND for HRS^51^.

***Health behaviors*** include smoker status as reported by respondents (never smoked, past smoker, and current smoker as the reference group), BMI split into five categories (underweight (less than 18.4), overweight (25 to 29.9), obese (30 to 34.9), morbidity obese (greater than or equal to 35), and normal weight (18.5 to 24.9) as the reference group), alcohol use based on number of drinks per day drinking (non-drinker, five or more drinks, and one to four drink as the reference group), and an index of physical activity (the sum of respondent reported light, moderate, and vigorous activity, each ranging from 1 (*never*) to 5 (*every day*)).

***Covariates.*** We control for batch and plate effects using batch (with the first batch as the reference group). Because percentages of blood cells change with age and blood cell composition can affect transcription levels, we control for blood cell composition using 7 genes indicative of cell type, *CD3D*, *CD19*, *CD4*, *CD8A*, *FCGR3A*, *NCAM1*, and *CD14*.

### Analytic Plan

Machine learning analyses were conducted in R 4.4.0 “Puppy Cup”^52^ using the tidyverse^53^, glmnet^54,55^, and lubridate^56^ packages. We restricted the set of genes used for training to coding genes with relatively high expression in human venous blood. Of the 50,611 genes that were measured and were successfully mapped in HRS, we restricted ourselves to the 19,291 protein coding genes and to genes with a mean count per million greater than 3 in the total HRS sample, leaving 10,964 genes.

We ran elastic net models using Cox regression to predict 4-year mortality hazard using these 10,964 genes, chronological age, and sex in a N = 1802 training set randomly selected from HRS^54,55^. Mortality hazard was assessed using month of death of participants known by HRS to have died. Hyperparameters, including the alpha and lambda penalty term, were selected using 5x cross validation with a grid search procedure. 11 alpha values were tested with more values close to 0 (viz., 0.000, 0.001, 0.008, 0.027, 0.064, 0.125, 0.216, 0.343, 0.512, 0.729, 1.000).

The alpha and lambda values that produced the lowest mean square error were selected. We adjusted the mean and variance to be the same as the mean and variance of HRS age in 2016 to make this an age-like variable.

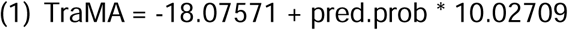

Where pred.prob is the log hazard of mortality from the Cox elastic net. We then tested this surrogate score in the N = 1794 testing set.

Validation in the HRS was conducted in R 4.4.0 “Puppy Cup”^52^ using the tidyverse^53^ and survey^57^ packages. Validation regressions and descriptive statistics in Table 1 and Figures 1 and 2 used survey weights provided by HRS for use in the RNAseq subsample (vbsi16wgtra). The total testing sample was 1794 participants. Because some participants were missing data on individual outcome variables and covariates, for mortality analyses N = 1791, for multimorbidity analyses N = 1791, for cognitive dysfunction N = 1791, and for ADLs and IADLs analyses N = 1588.

**Table 1b.** Descriptive Statistics for the LLFS dataset.

|  | Mean /<br>Proportion | SD |
| --- | --- | --- |
| Age | 70.04 | 15.6 |
| Subset: Proband | 0.32 |  |
| Subset: Offspring | 0.50 |  |
| Subset: Spouse | 0.18 |  |
| Gender: Female | 0.54 |  |
| Mortality | 0.22 |  |

Validation in the LLFS was conducted in R 4.4.0 “Puppy Cup”52 using coxme and dplyr packages. The total sample size was 1920 participants belonging to visit 1 in LLFS. TraMA was calculated using the algorithm developed in HRS. Mortality was analyzed using mixed-effect Cox proportional hazards regression models with adjustment for family effects.

Validation in public dataset was conducted in R 4.4.0 “Puppy Cup”^52^ using the tidyverse^53^ and MASS^58^ packages. For GSE171730, smoker status was analyzed using logistic regression, and pack years and FEV1 predicted percentage were assessed with linear regression. For GSE123658, diabetes status was assessed using logistic regression. For GSE186507, IBD status and active IBD status were assessed using logistic regression, and severity level was assessed using ordinal logistic regression. For GSE185263, sepsis status and mortality were assessed using logistic regression, and severity was assessed using linear regression.

## Data Availability

All data except LLFS are publicly available. LLFS data is available by request.

https://hrs.isr.umich.edu/about

https://longlifefamilystudy.com/

https://www.ncbi.nlm.nih.gov/geo/query/acc.cgi?acc=GSE171730

https://www.ncbi.nlm.nih.gov/geo/query/acc.cgi?acc=GSE123658

https://www.ncbi.nlm.nih.gov/geo/query/acc.cgi?acc=GSE186507

https://www.ncbi.nlm.nih.gov/geo/query/acc.cgi?acc=GSE185263

## Author Contributions

Eric T. Klopack (Conceptualization; Methodology; Formal analysis; Writing - Original Draft); Gokul Seshardri (Formal analysis; Validation; Writing - Review & Editing); Thalida Em Arpawong (Resources; Data Curation; Writing - Review & Editing); Steve Cole (Resources; Writing - Review & Editing); Bharat Thyagarajan (Resources; Data Curation; Writing - Review & Editing); Eileen M. Crimmins (Resources; Data Curation; Writing - Review & Editing; Supervision; Project administration; Funding acquisition)

---

**Figure S1.**
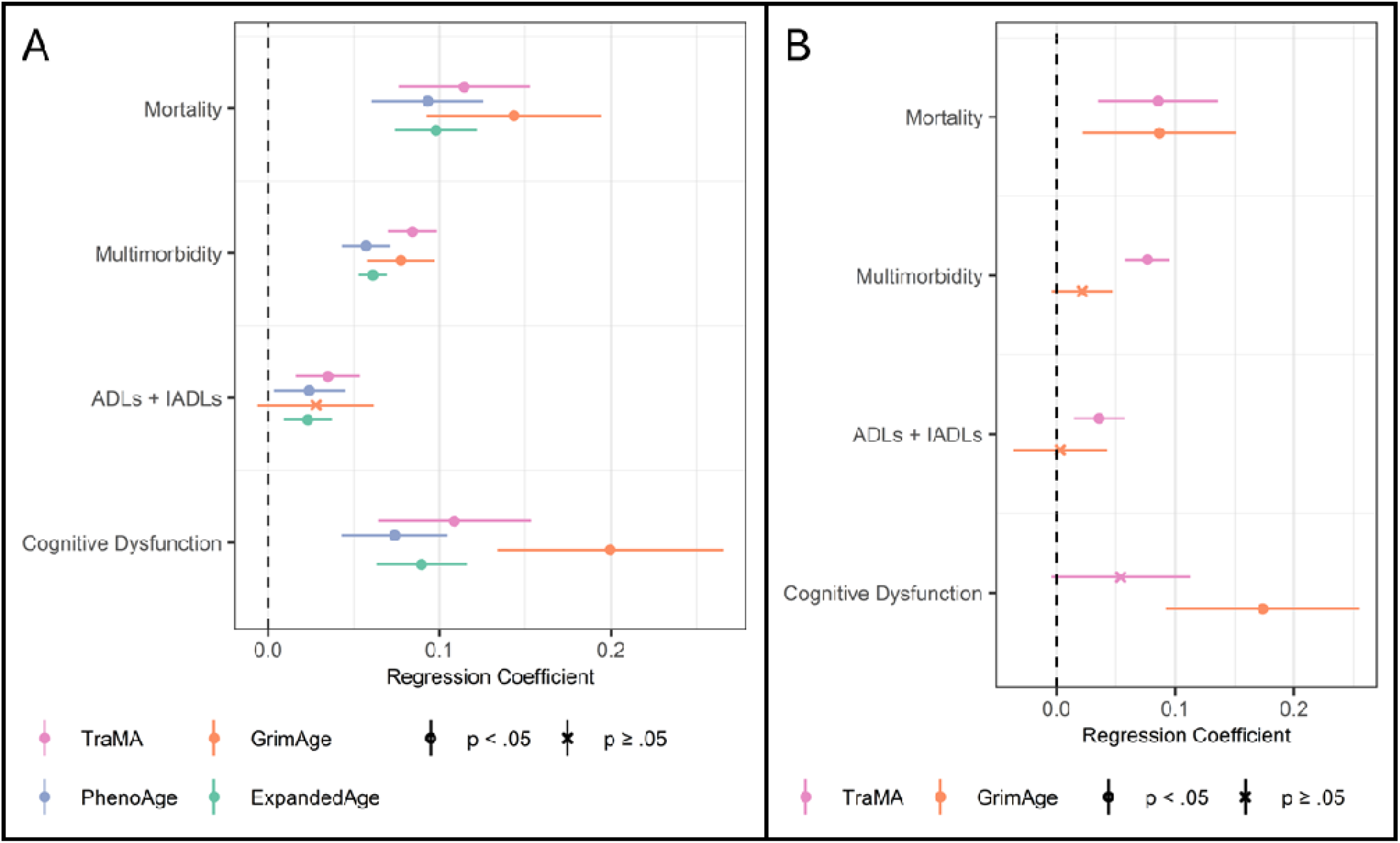
***Panel A.*** Regressions of health outcomes on biological aging measures; points represent regression coefficients and bars represent 95% confidence intervals; all models include age, race/ethnicity, sex/gender, and cell type as covariates; each point represents a separate regression equation. ***Panel B.*** Regressions of health outcomes on TraMA and GrimAge controlling for each other; points represent regression coefficients and bars represent 95% confidence intervals; all models include age, race/ethnicity, sex/gender, and cell type as covariates; each row represents a separate regression equation.

